# The Role of Sacubitril/Valsartan and Gliflozins in Heart Failure with Reduced Ejection Fraction after Cardiac Resynchronization Therapy

**DOI:** 10.1101/2023.03.30.23287985

**Authors:** Celeste Fonderico, Valerio Pergola, Daniele Faccenda, Gianluigi Comparone, Aldo Marrese, Alfonsomaria Salucci, Giuseppe Ammirati, Luigi Cocchiara, Alfonso Varriale, Giovanni Esposito, Antonio Rapacciuolo, Teresa Strisciuglio

## Abstract

**BACKGROUND:** The angiotensin receptor-neprilysin inhibitor (ARNi) and the sodium- glucose co-transporter 2 inhibitors (SGLT2i) have improved the outcome of patients with heart failure and reduced ejection fraction (HFrEF). However, data characterizing their effectiveness after cardiac resynchronization therapy (CRT) implant are relatively scarce. This study investigated the impact of ARNi and SGLT2i treatment 1) on CRT response at 12 months 2) on the cardiac function and the clinical functional status (NYHA class) at mid- and long-term follow-up 3) on the cardiac and overall survival at long-term follow-up.

**METHODS AND RESULTS:** HFrEF patients referred for CRT implant were enrolled in the study and were grouped by the ARNi/SGLT2i therapy. A first analysis investigated the synergistic impact of these drugs started at implant on 1-year CRT response and included all 172 patients enrolled. In order to evaluate whether the time of ARNi/SGLT2i initiation after CRT response assessment is meaningful, a second analysis considered 100 patients with a follow-up ≥ 24 months. The median follow-up was 63.1 (confidence interval [CI] 95%, 52.7 - 73.8) months.

At 1-year follow-up, 40 of 51 (78.4%) patients in ARNi or SGLT2i group and 66 of 121 (54.5%) in the no treatment group were classified as responders (p = 0.006). In multivariable analysis, ARNi/SGLT2i use was an independent predictor of CRT response (odds ratio, 5.38; CI 95%, 2-16.2; p = 0.001). At mid-term follow-up (median time [interquartile range, IQR] 40.6 [25.2; 58.3] months), 61 patients started to assume these drugs. NYHA functional class improved in 23 (37.7%) patients and decreased in only 2 (3.3%) in ARNi/SGLT2i patients vs 13 (33.3%) in no treatment group (p < 0.001). ARNi and SGLT2i improved significantly also the Δ LVEF, with a median [IQR] increase of 4 [2; 8] % compared to the no treatment group - 1.8 [-4; 0.2] % (p < 0.001) and were independently associated with a NYHA functional class II or I at long-term (hazard ratio [HR], 3.67; CI 95%, 1.37-10.2; p < 0.001). Their estimated effectiveness was consistent over the entire follow-up period (Schoenfeld residuals test, p = 0.10), although without reaching statistical significance effects on cardiovascular survival (HR, 0.61; CI 95%, 0.25-1.50; p = 0.22).

**CONCLUSIONS:** The ARNi and SGLT2i treatment in CRT patients improves the clinical and echocardiographic response at 12-month and long-term follow-up, independently from the time of initiation. These drugs also confer benefit on survival, however further studies are needed to confirm these data.

## INTRODUCTION

Heart failure (HF) is one of the greatest public health burdens worldwide ^1^. Since recent studies have demonstrated that angiotensin receptor-neprilysin inhibitor (ARNi or sacubitril/valsartan, S/V) and sodium-glucose co-transporter 2 inhibitors (SGLT2i or gliflozins) reduce the risk of cardiovascular mortality and worsening of heart failure in patients with reduced ejection fraction (HFrEF) ^2–4^, the current European guidelines recommend in class I as key first-line treatment the angiotensin-converting enzyme inhibitor (ACE-i) or ARNi and gliflozins on top of beta-blockers and mineralocorticoid receptor antagonist ^5^. Furthermore, the cardiac resynchronization therapy (CRT) is an established treatment for therapy-refractory mild to severe HFrEF patients with left ventricular conduction delay and is recommended for symptomatic patients despite optimal medical therapy for at least 3 months ^6^. The clinical benefit of ARNi and SGLT2i initiation is net in non-device-bearing patients ^7, 8^, however in large multicentric trials only a minority of patients were already implanted with a cardiac resynchronization therapy with defibrillator (CRTD), and in a real-world setting a significant gap in their prescription exists ^9^. Thus, it is less clear whether CRTD carriers may further benefit of the treatment with ARNi and SGLT2i even when starting them after the device implantation.

The present study aims to evaluate in a cohort of CRTD patients 1) the impact of ARNi and SGLT2i treatment on CRT response at 12 months after implantation 2) their impact at mid-term and long-term follow-up on cardiac function and clinical functional status (NYHA class) 3) the effects of these drugs on cardiac and overall survival 4) whether the time of initiation of these drugs influences the clinical response.

## METHODS

### Study population

This was a single-center observational retrospective study including all HFrEF symptomatic patients consecutively referred for CRTD implantation at the Department of Cardiology of Federico II University of Naples, from January 2015 to January 2022. All patients received CRTD according to the guidelines of the European Society of Cardiology ^10^ and were included in the local clinical database. Each patient signed the informed consent for data collection and for inclusion in the database.

In order to avoid possible confounding factors and make our population homogeneous, we included only ischemic (ICM) and non-ischemic (NICM) patients excluding other reversible causes of HF (such as acute viral myocarditis, alcohol-induced heart disease and tachycardia-related cardiomyopathy), valvular diseases ^11^, chemotherapy-induced cardiomyopathy ^12^ and dilated-phase hypertrophic cardiomyopathy ^13^. Other exclusion criteria were age below 18, lack of complete echocardiography or medical therapy data at 12-months follow-up and patients in ARNi or SGLT2i treatment > 3 months before CRTD implant.

Patients were divided into the following categories based on the pharmacological therapy at implantation time, and subsequently after CRT response assessment: 1) not in ARNi and SGLT2i 2) in ARNi and/or SGLT2i treatment.

The study protocol conforms to the ethical guidelines of the 1975 Declaration of Helsinki and was approved by the Institutional Ethics Committee.

### Baseline characteristics and follow-up data

Data retrieved from the local database included demographic variables, cardiovascular risk factors, pharmacological therapy, underlying heart disease, New York Heart Association (NYHA) functional class, echo- and electrocardiographic parameters.

Following implantation, patients underwent ambulatory visits scheduled at 1, 6 and 12 months. After 12 months, the time of visit and of echocardiography was at the discretion of the care provider. Device interrogation was performed within 2 months from the implant and at 12 months, with evaluation of biventricular pacing and optimization protocol. Any adjustment in therapy, particularly for ARNi and gliflozins use, was recorded. For the analysis, we considered the last dose of ARNi if changed < 2 months from the CRT implant, otherwise the dose taken for most of the time during the first year. 3 patients discontinued ARNi <1 month after CRT due to side effects and were considered in the no ARNi/SGLT2i group.

During follow-up visits, clinical data (including class NYHA) were collected and all clinical events, including HF and non-cardiac rehospitalizations, were recorded. Data on overall mortality were assessed on the basis of hospital records or from telephonic interviews with caregivers.

### Echocardiography Data and Heart Failure etiology definition

A standard echocardiographic examination was performed in all patients prior to CRT implantation and at least twice in the first year of follow-up. Chamber quantification was realized conforming to current recommendations. LV volume was assessed using Simpson’s biplane method and indexed to body surface area. Ejection Fraction was calculated from LV volumes according to clinical practice guidelines.

Pre-CRT, 12 months after device therapy and long-term (until the last follow-up) echocardiography data were collected by expert cardiologist. Left ventricular structural and functional alterations were assessed by absolute change in left ventricular ejection fraction (LVEF) and percentage change in left ventricular end-systolic volume (LVESV).

Before CRTD implantation, all patients without a known coronary artery assessment underwent to coronarography at our center. Patients were classified as ICM if they had a documented history of myocardial infarction or of a coronary revascularization procedure (prior coronary artery bypass graft surgeries or percutaneous balloon and/or stent angioplasty) or a significant coronary artery disease at coronarography with angina pectoris or other coronary-related symptoms. NICM was defined in the absence of each of the above criteria of ICM, implying a systolic dysfunction leading to HF not due to coronary disease or other recognized cause (such us primary valvular disease, reversible cause of HF, hypertrophic cardiomyopathy).

### Definition of clinical and echocardiographic response to CRT

Clinical response was evaluated at 12 months after CRT implant using the Clinical Response (CR) definition ^14^, considering a hierarchical composite criterion comprising live status, HF hospitalization occurrence and variation in NYHA functional class. In details, a positive response was assigned to patients who remained alive without any HF hospitalization during the first year and experienced an improvement of almost 1 NYHA class or remained in NYHA class I or II. On the other hand, patients who died or were hospitalized for HF or showed worsening of their NYHA class or not improvement from NYHA class III and IV were classified as non-responders.

Furthermore, patients were classified as echocardiographic CRT responders if they had LV reverse remodeling at 12 months, evaluated as a LVEF improvement > 5% or a LVESV reduction > 15%.

### End Points

In the first analysis (Figure 2), patients were divided in two groups, ARNi/SGLT2i group and no ARNi/SGLT2i group, according to whether they started or not the drugs at the time of CRT implant. The primary endpoint of this analysis was the evaluation of the clinical (CR and class NYHA) and echocardiographic response to CRT at 12 months. The second analysis included patients who initiated these drugs ≥12 months after the CRT implant (mid-term initiation ARNi/SGLT2i group) and with a minimum follow-up of 24 months. For those patients who never started these drugs (no ARNi/SGLT2i group), the mid-term follow-up was considered as the visit closest to the median follow-up of the ARNi/SGLT2i group. The primary endpoint of the second analysis was the clinical and echocardiographic response at long-term follow-up (last available visit) and the secondary endpoints were the composite of total cardiovascular deaths and separately cardiac or non-cardiac deaths. The patients were censored at the outcome events or at the end of the follow-up period (Jan 2023). Importantly, patients included in this second analysis had a follow-up > 6 months after drugs prescription, otherwise were considered in the no ARNi/SGLT2i group.

The study protocol is summarized in Figure S1 (in the Supplement).

### Statistical Analysis

Demographic and clinical data referred to the baseline were summarized using standard descriptive statistics. Data distribution was assessed through visual analysis of the boxplot for each variable. Continuous variables were expressed as mean ± Standard Deviation (SE) if normally distributed or as median [inter-quartile range (IQR)] in the case of skewed distribution and compared between groups by means of a t-test for unpaired samples or Wilcoxon-Mann-Whitney non-parametric test, respectively; categorical variables were reported as absolute and relative frequencies and comparisons between groups were performed with χ2 test. Pairwise testing with the Holm correction was applied to account for multiple comparisons ^15^. The Analysis of Variance (one-way ANOVA) was used to test all factorial covariates with more than two levels.

For the primary outcome, to identify the potential predictors of clinical response to CRT, we used a stepwise logistic regression: first we applied a univariable logistic regression model to test the relationship between our primary endpoint (dependent variable) and all the clinical findings, including the use of ARNi and SGLT2i (independent variable). Characteristics significantly (p < 0.05) or nearly significantly (p < 0.10) associated with the outcome in the univariable analysis were first entered as candidate variables in a multivariable logistic regression analysis. The included independent variables were tested for collinearity to exclude possible confounders. The final multivariable model was selected using a backward-elimination algorithm after testing residual deviance with ANOVA. In a similar way, we ran a multivariable linear regression model to evaluate the correlation between the covariates and the change in LVEF. Results of these models were expressed as odds ratios (ORs) and mean differences with the corresponding 95% confidence intervals (CIs).

Regarding the co-primary outcome, to assess the clinical main endpoint of NYHA class improvement or remained in I or II and to solve the possible differences in time between mid- and long-term follow-up, a multivariable Cox proportional hazard regression model was performed by using hazard ratios (HRs) and 95 % CIs. In addition, to test the assumption of proportional hazard particularly regarding the ARNi/SGLT2i treatment, we constructed another Cox regression model considering the follow-up time from CRT implant and used the quantitative test of Schoenfeld residuals, with a p value < 0.05 allowed to reject the null hypothesis of proportionality; we finally plotted the graphic shown the Schoenfeld residuals and the ARNi/SGLT2i-related coefficient arising from the multivariable Cox regression model and accounting the entire follow-up period ^16^.

To analyze the impact of ARNi, SGLT2i or both treatments and the different ARNi doses tested individually, we limited only to a descriptive analysis due to the low sample size after dividing in the subgroups, without inferential statistics.

Furthermore, in order to analyze the risk of experiencing the secondary outcomes, we used Kaplan-Meier curves to estimate the outcomes-free survival function stratified by assumption of S/V and gliflozins. Finally, the Cox proportional hazards regression analysis was used to quantify the association with the main covariates. Analogous to the previous description, from a univariable model we constructed the multivariable analysis with a stepwise selection.

Importantly, in all Cox regression analysis ARNi and SGLT2i treatment was considered a time-dependent covariate to account for patients who stopped or started the therapy during the follow-up ^17^. We also tested the linearity (for continuous variables) and proportional hazard (for all the covariates) assumptions computing restricted cubic spline bases and Schoenfeld residuals, respectively, to assess the best fit of all proportional hazard analysis ^16, 18, 19^.

Clinically relevant interactions with the main covariate were tested in all models.

All tests were two tailed, and values of p < 0.05 were considered significant. Statistical analysis was performed using R version 4.2.1 (R foundation, Vienna, Austria).

The data that support the findings of this study are available from the corresponding author upon reasonable request.

## RESULTS

### Study population

Overall, 240 patients underwent CRTD implant, of these 172 were included in our study population (Figure 1). During the first-year, 51 (29.7 %) patients were assuming ARNi and/or SGLT2i treatment (ARNi/SGLT2i group) and 121 (70.3 %) weren’t (No ARNi/SGLT2i group). In the ARNi/SGLT2i group, timing and doses of these drugs varied among patients as evidenced in supplementary Figure S2.

**FIGURE 1.**
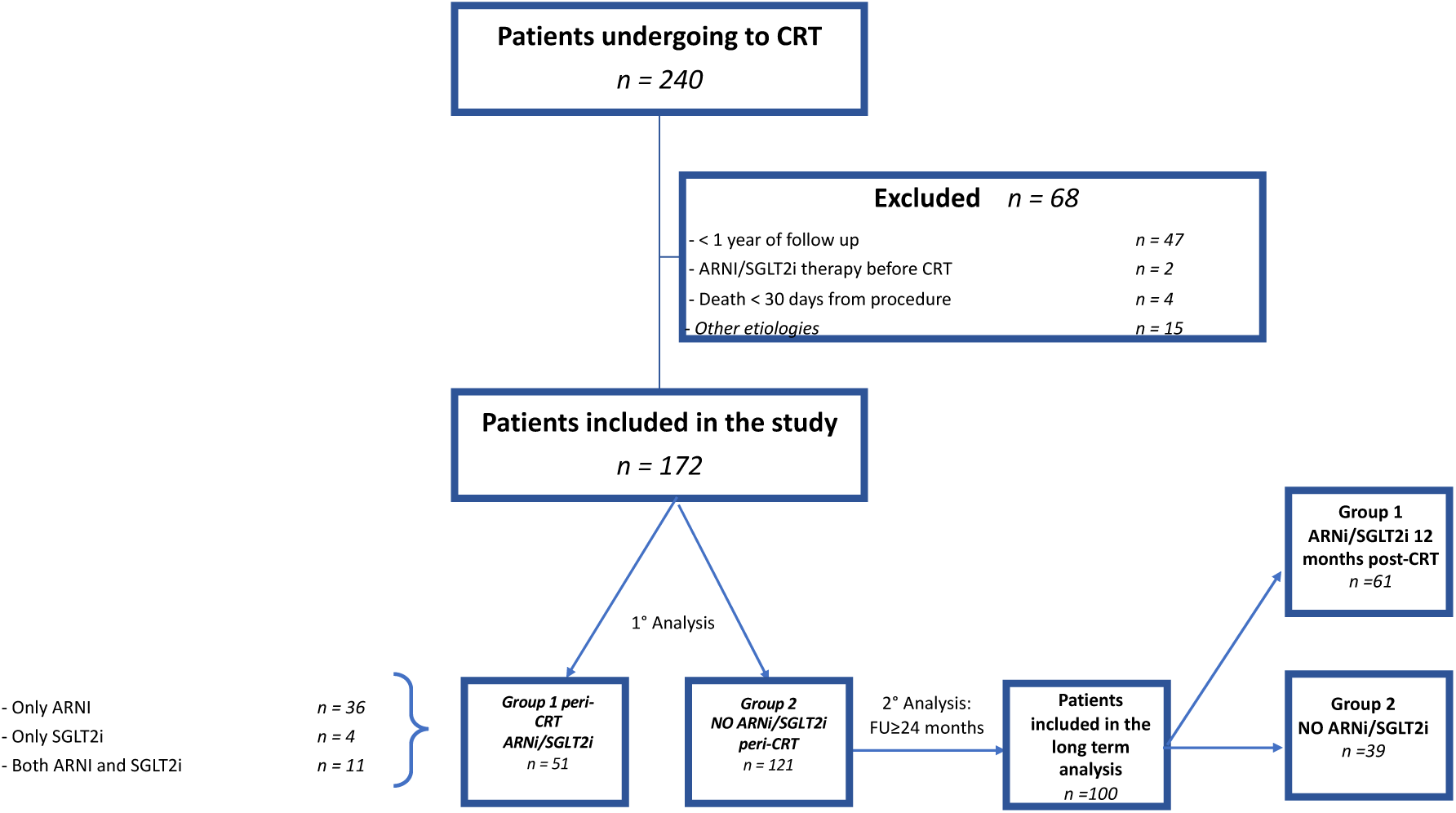
Flow chart of the recruitment process: 240 consecutive CRT-D patients were initially enrolled meeting the inclusion criteria of HF diagnosis with EF ≤ 35%. Of these, 42 (19.6 %) were missing 1-year clinical or echocardiogram follow-up, 5 due to cardiovascular implantable electronic device infections requiring extraction, 15 (6.3 %) had other HF etiology, 2 (0.8 %) were already in treatment with ARNi (5 and 10 months before implant procedures) and an additional 3 (1.2 %) patients died < 3 months after implantation (none of theme was in ARNi/SGLT2i treatment) and were all excluded.

**FIGURE 2.**
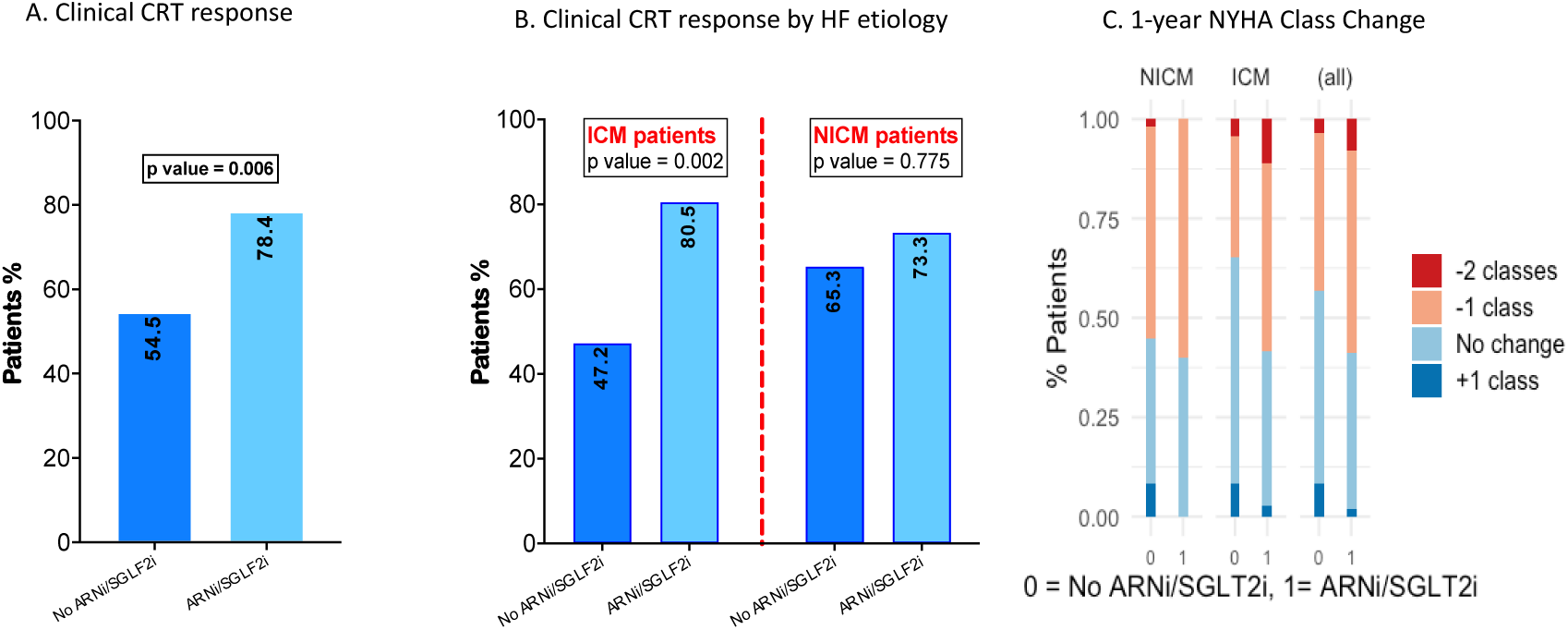
Clinical response to CRT and functional status at 1-year. One-year clinical response to CRT in no ARNi/SGLT2i and ARNi/SGLT2i groups in overall population **(A)** and based on HF etiology **(B).** Effectiveness of ARNi and SGLT2i treatment in NYHA functional class change at 1-year follow-up compared to baseline in overall population and dividing by HF etiology **(C)**. NICM refers to Non-Ischemic Cardiomyopathy, ICM to Ischemic Cardiomyopathy patients. Statistics: χ2 test.

Baseline characteristics are shown in Table 1. The baseline QRS duration was similar (157.5 (± 8.9) vs 158 (± 12.9), p = 0.303) with most patients in sinus rhythm at implantation (79.3 % vs 88.3 %, p = 0.242). Most patients were on optimal medical therapy, with a large proportion treated with a renin–angiotensin–aldosterone system inhibitor and a β-blocker. However, only the 45% and 46% were taking β-blocker at the target dose in ARNi/SGLT2i group and in the no ARNi/SGLT2i group, respectively.

**TABLE 1.** Clinical characteristics of the overall population and dividing by ARNi and SGLT2i treatments. Baseline characteristics and treatments in overall population (n =172) and dividing by no ARNi/SGLT2i (n = 121) and ARNi/SGLT2i (n =51) groups. Data are expressed as number (%), mean ± standard deviation or median (25th; 75th percentile). In bold are reported significant values. Biventricular pacing refers to the 1-year finding. Glomerular filtration rate was calculated in ml/min/1.73m^2^, baseline QRS duration in milliseconds. NICM = Non-Ischemic Cardiomyopathy; ICM = Ischemic Cardiomyopathy; COPD = Chronic Obstructive Pulmonary Disease; TIA = Transient Ischemic Attack; LVESV = Left Ventricular End-Systolic volume; LVEF = Left Ventricular Ejection Fraction; ACE-I/ARB = Angiotensin Converting Enzyme Inhibitors/Angiotensin II Receptor Blockers; MRA = Mineralocorticoid Receptor Antagonist. Statistics: t-student, Wilcoxon-Mann-Whitney and χ2 tests; one-way ANOVA.

| **Variable** | **Overall population** | **No ARNi/SGLT2i** | **ARNi/SGLT2i** | **p value** |
| --- | --- | --- | --- | --- |
| Age - years | 67.7 [60.9; 75.7] | 67.2 [59.7; 75] | 69.1 [63.3; 76.5] | 0.334 |
| Male sex - no. (%) | 138 (80.2) | 93 (76.9) | 45 (88.2) | 0.133 |
| CRT-D upgrade - no. (%) | 40 (23.2) | 28 (22.6) | 12 (23.5) | 1 |
| Etiology - no. (%) |  |  |  | 0.229 |
| NICM | 64 (37.2) | 49 (40.5) | 15 (29.4) |  |
| ICM | 108 (62.7) | 72 (59.5) | 36 (70.6) |  |
| NYHA Class - no. (%) |  |  |  | 0.621 |
| II | 50 (28.6) | 33 (26.6) | 17 (33.3) |  |
| III | 108 (62.8) | 77 (63.6) | 31 (60.8) |  |
| IV | 14 (8.1) | 11 (9.1) | 3 (5.9) |  |
| **Cardiac risk factors - no. (%)** |  |  |  |  |
| Treatment for Hypertension | 145 (84.3) | 100 (82.6) | 45 (88.2) | 0.489 |
| Atrial Fibrillation | 56 (32.6) | 40 (33.1) | 16 (31.4) | 0.970 |
| Dyslipidemia | 118 (68.6) | 78 (64.5) | 40 (78.4) | 0.105 |
| Diabetes Mellitus | 76 (44.2) | 54 (44.6) | 22 (43.1) | 0.991 |
| **Other Comorbidities** |  |  |  |  |
| COPD - no. (%) | 51 (29.7) | 41 (33.9) | 10 (19.6) | **0.025** |
| Glomerular filtration rate | 61.2 (±24.1) | 59.6 (± 25.5) | 64.9 (± 20.2) | 0.150 |
| Previous stroke or TIA - no. | 14 (8) | 8 (6.6) | 6 (11.8) | 0.410 |
| **Echocardiographic findings** |  |  |  |  |
| LVESV - ml | 187 (± 43) | 188 (±69) | 181 (± 77) | 0.124 |
| LVEF - % | 27.7 (± 4.9) | 27.4 (± 5.1) | 28.7 (± 4.5) | 0.104 |
| **Electrocardiographic findings** |  |  |  |  |
| Left bundle-branch block - no. | 116 (67.4) | 82 (67.7) | 34 (66.6) | 0.278 |
| Baseline QRS duration | 156.1 (± 10.4) | 157.5 (± 8.9) | 158 (± 12.9) | 0.303 |
| Sinus rhythm at implantation - | 141 (81.9) | 96 (79.3) | 45 (88.3) | 0.242 |
| Biventricular pacing, % | 97.5 [96; 99] | 97 [96; 99] | 98 [96; 100] | 0.206 |
| **Medications - no. (%)** |  |  |  |  |
| Beta-blocker | 159 (92.4) | 113 (93.4) | 46 (90.2) | 0.651 |
| ACEi/ARB (excluding ARNI) | 114 (66.3) | 112 (92.5) | 2 (3.9) | - |
| Amiodarone | 24 (13.9) | 13 (10.7) | 11 (21.6) | 0.103 |
| Loop Diuretic | 144 (83.7) | 107 (88.4) | 37 (72) | **0.019** |
| MRA | 87 (50.5) | 65 (53.7) | 22 (43.1) | 0.271 |
| Ivabradine | 6 (3.5) | 5 (4.1) | 1 (1.9) | 0.799 |
| Digitalis | 5 (2.9) | 3 (2.4) | 2 (3.9) | 0.921 |
| Lipid-lowering treatment | 142 (82.5) | 97 (80.2) | 45 (88.2) | 0.225 |
| Anticoagulants | 56 (32.6) | 39 (32.2) | 17 (33.3) | 1 |
| ARNi - no. (%) | - | - | 47 (92.2) | na |
| SGLT2i - no. (%) | - | - | 15 (29.4) | na |
| ARNi and SGLT2i - no. (%) | - | - | 11 (21.5) | na |

Compared with the no ARNi/SGLT2i group, patients in ARNi/SLGT2i treatment were less likely to have chronic obstructive pulmonary disease (COPD) (19.6 % vs 33.9 %, p = 0.025) and Loop Diuretic medication (72 % vs 88.4 %, p = 0.019).

### Twelve-months clinical response to CRT

During the first 12-month follow-up, 10 patients (5.8 %) died, all because of acute heart failure decompensation and were considered not clinical responder. Of these, only 1 patient was in therapy with low dose of ARNi (male, NYHA class IV, suffered from ICM).

Based on the CR definition, 78.4 % of patients in the ARNi/SGLT2i and 54.5 % of patients in the no ARNi/SGLT2i group were classified as responders (P = 0.006, Figure 2A). Univariable logistic regression tests disclosed a significant relationship between ARNi/SGLT2i treatment and clinical response [p = 0.04, OR 3.03 (CI 1.46 - 6.71)] (Table 2). In the multivariable analysis, ARNi/SGLT2i use was confirmed as a strong predictor [p = 0.001, OR 5.38 (CI 2 – 16.2)]. History of atrial fibrillation, a lower biventricular pacing percentage and right bundle branch block (RBBB) patients were negatively associated with the outcome.

**TABLE 2.**
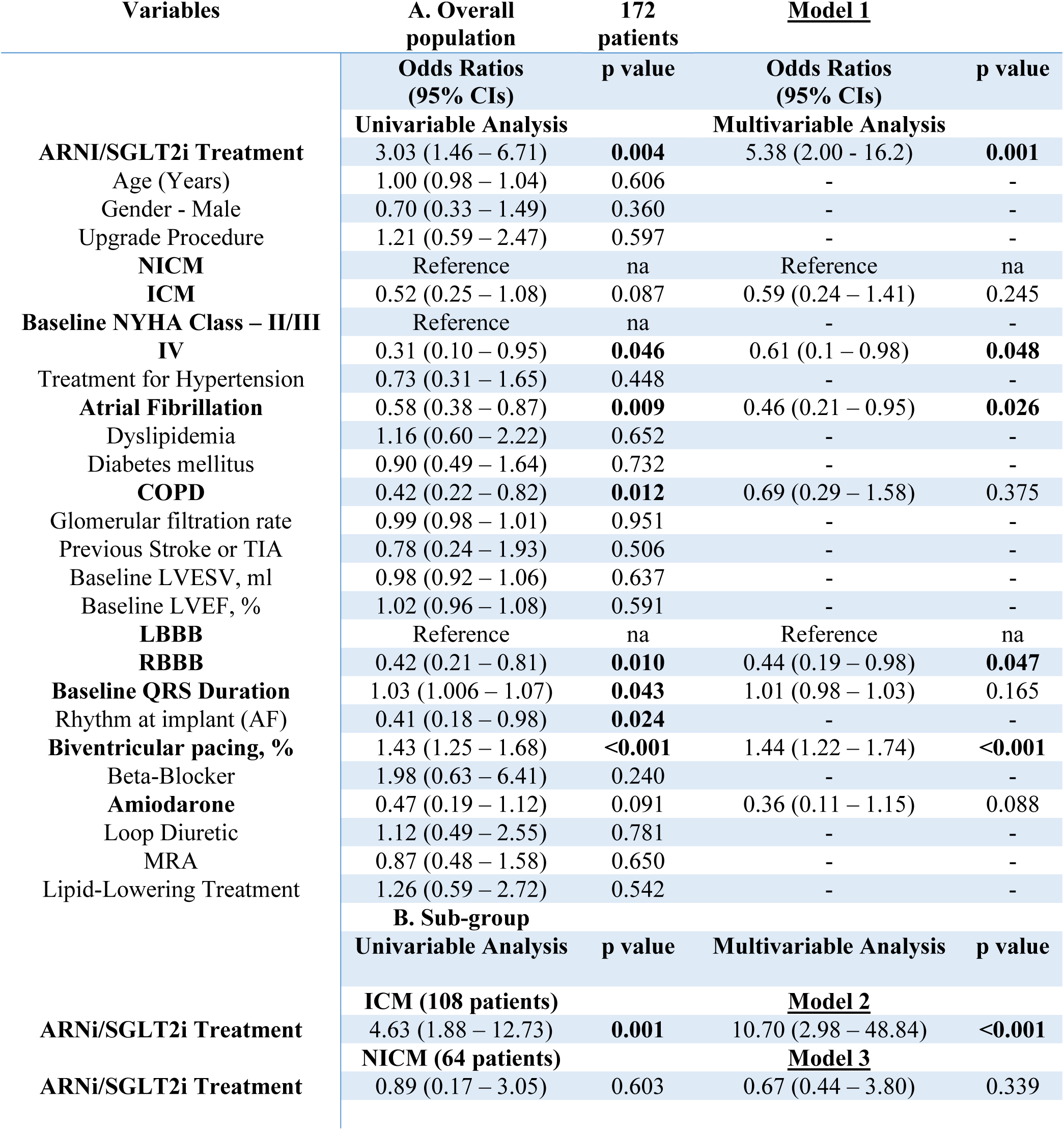
Predictors of clinical CRT response in overall population and divided by HF etiology. Predictors of clinical CRT response based on logistic regression analysis, **A.** Univariable and multivariable (Model 1) logistic regression analysis in overall population. In bold are reported significant values and the covariates entered in the multivariable analysis. After multiple logistic regression model testing residual deviance with ANOVA, we excluded from the multivariable analysis the covariate Sinus Rhythm at implant due to correlation with Atrial Fibrillation. Only one subgroup interaction was identified, between the main covariate ARNi/SGLT2i treatment and heart failure etiology (p = 0.028). All other interactions exceeded 0.10. **B.** Univariable and multivariable logistic regression analysis dividing by heart failure etiology. Model 2 and Model 3 were adjusted for the same covariates of Model 1, except for Heart failure Etiology. Biventricular pacing rate refers to the 1-year finding. Glomerular filtration rate was calculated in ml/min/1.73 m2, baseline QRS duration in milliseconds. NICM = Non-Ischemic Cardiomyopathy; ICM = Ischemic Cardiomyopathy; COPD = Chronic Obstructive Pulmonary Disease; TIA = Transient Ischemic Attack; LVESV = Left Ventricular End-Systolic volume, in milliliters; LVEF = Left Ventricular Ejection Fraction; LBBB = Left Bundle-Branch Block; RBBB = Right Bundle-Branch Block; AF = Atrial fibrillation; MRA = Mineralocorticoid Receptor Antagonist.

| Variables | A. Overall population | 172 patients | <u>Model 1</u> |  |
| --- | --- | --- | --- | --- |
|  | Odds Ratios (95% CIs) | p value | Odds Ratios (95% CIs) | p value |
|  | **Univariable Analysis** |  | **Multivariable Analysis** |  |
| **ARNI/SGLT2i Treatment** | 3.03 (1.46 – 6.71) | **0.004** | 5.38 (2.00 - 16.2) | **0.001** |
| Age (Years) | 1.00 (0.98 – 1.04) | 0.606 | - | - |
| Gender - Male | 0.70 (0.33 – 1.49) | 0.360 | - | - |
| Upgrade Procedure | 1.21 (0.59 – 2.47) | 0.597 | - | - |
| **NICM** | Reference | na | Reference | na |
| **ICM** | 0.52 (0.25 – 1.08) | 0.087 | 0.59 (0.24 – 1.41) | 0.245 |
| **Baseline NYHA Class – II/III** | Reference | na | - | - |
| **IV** | 0.31 (0.10 – 0.95) | **0.046** | 0.61 (0.1 – 0.98) | **0.048** |
| Treatment for Hypertension | 0.73 (0.31 – 1.65) | 0.448 | - | - |
| **Atrial Fibrillation** | 0.58 (0.38 – 0.87) | **0.009** | 0.46 (0.21 – 0.95) | **0.026** |
| Dyslipidemia | 1.16 (0.60 – 2.22) | 0.652 | - | - |
| Diabetes mellitus | 0.90 (0.49 – 1.64) | 0.732 | - | - |
| **COPD** | 0.42 (0.22 – 0.82) | **0.012** | 0.69 (0.29 – 1.58) | 0.375 |
| Glomerular filtration rate | 0.99 (0.98 – 1.01) | 0.951 | - | - |
| Previous Stroke or TIA | 0.78 (0.24 – 1.93) | 0.506 | - | - |
| Baseline LVESV, ml | 0.98 (0.92 – 1.06) | 0.637 | - | - |
| Baseline LVEF, % | 1.02 (0.96 – 1.08) | 0.591 | - | - |
| **LBBB** | Reference | na | Reference | na |
| **RBBB** | 0.42 (0.21 – 0.81) | **0.010** | 0.44 (0.19 – 0.98) | **0.047** |
| **Baseline QRS Duration** | 1.03 (1.006 – 1.07) | **0.043** | 1.01 (0.98 – 1.03) | 0.165 |
| Rhythm at implant (AF) | 0.41 (0.18 – 0.98) | **0.024** | - | - |
| **Biventricular pacing, %** | 1.43 (1.25 – 1.68) | **<0.001** | 1.44 (1.22 – 1.74) | **<0.001** |
| Beta-Blocker | 1.98 (0.63 – 6.41) | 0.240 | - | - |
| **Amiodarone** | 0.47 (0.19 – 1.12) | 0.091 | 0.36 (0.11 – 1.15) | 0.088 |
| Loop Diuretic | 1.12 (0.49 – 2.55) | 0.781 | - | - |
| MRA | 0.87 (0.48 – 1.58) | 0.650 | - | - |
| Lipid-Lowering Treatment | 1.26 (0.59 – 2.72) | 0.542 | - | - |
|  | **B. Sub-group** |  |  |  |
|  | **Univariable Analysis** | **p value** | **Multivariable Analysis** | **p value** |
|  | **ICM (108 patients)** |  | **Model 2** |  |
| **ARNi/SGLT2i Treatment** | 4.63 (1.88 – 12.73) | **0.001** | 10.70 (2.98 – 48.84) | **<0.001** |
|  | **NICM (64 patients)** |  | **Model 3** |  |
| **ARNi/SGLT2i Treatment** | 0.89 (0.17 – 3.05) | 0.603 | 0.67 (0.44 – 3.80) | 0.339 |

Importantly, the only significant interaction in the multivariable model was between ARNi/SGLT2i treatment and heart failure etiology (p of interaction = 0.028). In the subsequent analysis of sub-groups divided according to heart failure etiology, we observed that only in ICM patients ARNi/SGLT2i use was a strong predictor of clinical response [p < 0.001, OR 10.7 (CI 2.98 – 48.84)] (Table 2, model 2 and 3).

Considering ICM patients, 29 of 36 (80.5 %) in the ARNi/SGLT2i group vs 34 of 72 (47.2 %) in the no ARNi/SGLT2i group were considered clinical responder (p = 0.002), against 11 of 15 (73.3 %)) vs 32 of 49 (65.3 %) in NICM patients (p = 0.775, Figure 2B).

As for the NYHA functional class, it decreased by at least 1 class in the ARNi/SGLT2i vs No ARNi/SGLT2i group in 60 % vs 42.8 % (P = 0,041, Figure 2C), and decreased by 2 classes in 8 % vs 2.67 %, P = 0.237). Once again, a greater benefit of the ARNi/SGLT2i therapy was significantly confirmed only in the ICM group: 60 % vs 33.3 % (P = 0,033, Figure 2C). The only 2 patients in NYHA class IV in the ARNi/SGLT2i group did not improve their clinical status.

Figure S3 in the supplement details the CRT response rate and NYHA functional class change according to ARNi, SGLT2i or both treatment and ARNi doses. Clinical benefits were found in all treatment groups, slightly more in both ARNi and SGLT2i recipients, whereas the 24/26 mg ARNi dose use appeared to have a lower effectiveness.

### Twelve-months echocardiographic CRT response

At 12-months, there were more CRT responders in the ARNi/SGLT2i group than in the no ARNi/SGLT2i group (76 % vs 50 %, respectively; p = 0.003, Fig. 3A). Table S1 in the supplement materials details the predictors of echocardiographic CRT response based on univariable and multivariable logistic regression analysis. Although the interaction between HF etiology and ARNi/SGLT2i treatment was not significant (p = 0.263), in NICM 9 of 11 ARNi/SGLT2i patients (81 %) and 29 of 50 (58 %) in the other group were considered CRT responders, without reaching statistical significance (p = 0.118).

**FIGURE 3.**
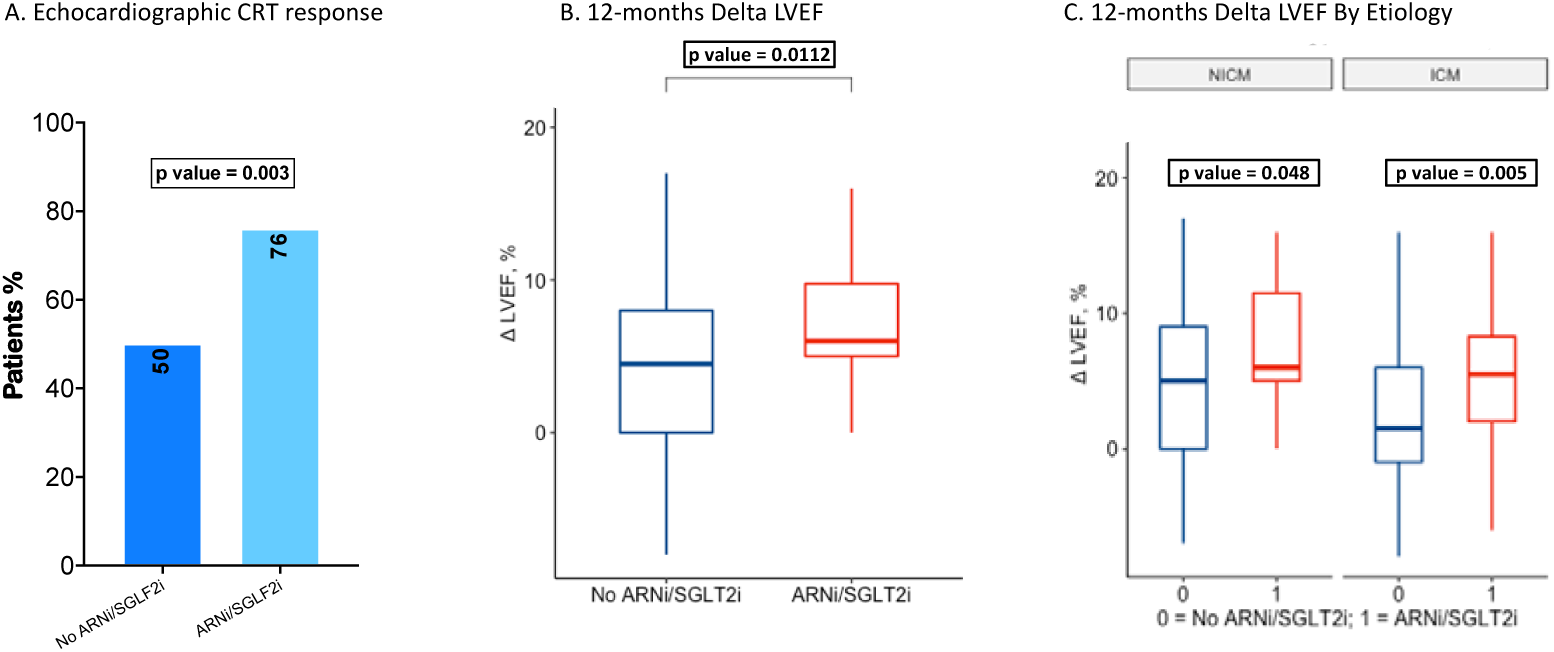
Echocardiographic assessment at 1-year follow-up. One-year rates of echocardiographic CRT response (evaluated as an improvement of 5% of LVEF or a reduction of 15% of LVESV) in 162 alive patients according to ARNi and SGLT2i treatment **(A).** Impact of ARNi and SGLT2i therapy on Delta LVEF change at 12-month echocardiographic evaluation in overall 162 patients **(B)** and dividing by HF etiology **(C)**. Boxplots show the median (central mark) with the 25^th^ and 75^th^ percentiles (box edges), and minimum and maximum values (whiskers). NICM refers to Non-Ischemic Cardiomyopathy, ICM to Ischemic Cardiomyopathy patients. Statistics: χ2 and Wilcoxon-Mann-Whitney tests.

The Δ LVEF increased of 6 % [IQR 5; 9.75 %] in ARNi/SGLT2i group vs 4.5 % [IQR 0 %; 8 %] in the no ARNi/SGLT2i group (p = 0.0112) (Fig. 3B).

Based on the linear regression analysis (Table 3), ARNi/SGLT2i treatment was significantly associated with a Δ LVEF 2.43 % average increase higher than no ARNi/SGLT2i group [p = 0.018, Estimate Coefficient = 2.43 (0.41 – 4.44)], confirmed in the multivariable model [p = 0.007, Estimate Coefficient = 2.5 (0.69 – 4.30)]. Indeed, COPD, RBBB and atrial fibrillation remained significant negatively and adequate biventricular pacing positively associated with Δ LVEF improvement; instead, ICM was not a negative predictor of Δ LVEF increase.

**TABLE 3.** Univariable and multivariable linear regression analysis assessed predictors associated with 12-month Δ LVEF improvement. Linear regression analysis assessed predictors associated with 12-month Δ LVEF improvement in 162 patients alive at 1-year follow-up. In bold are reported significant values and the covariates entered in the multivariable analysis. The covariate Sinus Rhythm at implant was excluded from the multivariable due to correlation with Atrial fibrillation. All interactions were not significant. Biventricular pacing refers to the 1-year finding. Glomerular filtration rate was calculated in ml/min/1.73 m^2^, baseline QRS duration in milliseconds. NICM = Non-Ischemic Cardiomyopathy; ICM = Ischemic Cardiomyopathy; COPD = Chronic Obstructive Pulmonary Disease; TIA = Transient Ischemic Attack; LVESV = Left Ventricular End-Systolic volume, in milliliters; LVEF = Left Ventricular Ejection Fraction; LBBB = Left Bundle-Branch Block; RBBB = Right Bundle-Branch Block; AF = Atrial fibrillation; MRA = Mineralocorticoid Receptor Antagonist.

| Variables | $\beta$ Regression Coefficients (95% CIs) | p value | $\beta$ Regression Coefficients (95% CIs) | p value |
| --- | --- | --- | --- | --- |
|  | Univariable Analysis |  | Multivariable Analysis |  |
| **ARNI/SGLT2i Treatment** | 2.43 (0.41; 4.44) | **0.0185** | 2.5 (0.69; 4.30) | **0.007** |
| Age (Years) | 0.02 (-0.06; 0.11) | 0.592 | - | - |
| Gender - Male | 0.80 (-1.54; 3.14) | 0.502 | - | - |
| Upgrade Procedure | 0.92 (-1.35; 3.19) | 0.425 | - | - |
| **NICM** | Reference | na | Reference | na |
| **ICM** | -1.59 (-3.53; 0.33) | 0.10 | -1.16 (-2.89; 0.57) | 0.186 |
| Baseline NYHA Class II | Reference | na | - | - |
| III | -1.36 (-3.43; 0.69) | 0.193 |  |  |
| IV | -1.71 (-5.8; 2.43) | 0.415 |  |  |
| Treatment for Hypertension | 0.62 (-1.95; 3.20) | 0.633 | - | - |
| **Atrial Fibrillation** | -1.73 (-2.98; -0.47) | **0.007** | -0.98 (-2.13; 0.160) | 0.090 |
| Dyslipidemia | 0.66 (-1.37; 2.69) | 0.522 | - | - |
| Diabetes mellitus | -0.36 (-2.26; 1.54) | 0.709 | - | - |
| **COPD** | -2.72 (-4.78; -0.67) | **0.009** | -1.92 (-3.77; -0.06) | **<0.001** |
| Glomerular filtration rate | -0.01 (-0.05; 0.03) | 0.710 | - | - |
| Previous Stroke or TIA | 0.70 (-2.62; 4.03) | 0.675 | - | - |
| Baseline LVESV, ml | 0.09 (-0.04; 0.12) | 0.460 | - | - |
| Baseline LVEF, % | -0.03 (-0.23; 0.125) | 0.712 | - | - |
| **LBBB** | Reference | na | Reference | na |
| **RBBB** | -3.66 (-5.62; -1.71) | **<0.001** | -3.67 (-5.44; -1.90) | **<0.001** |
| **Baseline QRS Duration** | 0.10 (0.01; 0.19) | **0.024** | 0.02 (-0.05; 0.10) | 0.530 |
| Rhythm at implant- AF | -2.22 (-4.78; 0.32) | 0.08 | - | - |
| **Biventricular pacing, %** | 0.47 (0.30; 0.63) | **<0.001** | 0.37 (0.20; 0.54) | **<0.001** |
| Beta-Blocker | 0.96 (-2.64; 5.57) | 0.598 | - | - |
| Amiodarone | -0.75 (-3.51; 2.01) | 0.59 | - | - |
| Loop Diuretic | -0.24 (-2.77; 2.30) | 0.854 | - | - |
| MRA | -0.74 (-2.63; 1.15) | 0.441 | - | - |
| Lipid-Lowering Treatment | 0.42 (-1.89; 2.72) | 0.772 | - | - |

Specifically, in the ARNi/SGLT2i group compared to no ARNi/SGLT2i group, Δ LVEF improved in both NICM [6.5 % (IQR 5; 11.5 %) vs 5 % (IQR 0; 8.5 %); p= 0.048] and ICM patients [5.5 % (IQR 2 %; 8.25 %) vs 1.5 % (IQR -1; 6 %), p = 0.005, Figure 3C].

The supplementary Figure S4 details the percentages of patients with different 1-year LVEF changes based on ARNi/SGLT2i treatment. Among non-responders, less patients in the ARNi/SGLT2i group than in the no ARNi/SGLT2i group had no increase or a reduction of LVEF (Δ LVEF <0%) (6 % vs 24.1 %, p = 0.004). No significant differences were found among groups in the rate of super-responders (LVEF improvement ≥ 10%).

### Impact of ARNi/SGLT2i on long-term follow-up

Overall, 94 patients were not taking ARNi and/or SGLT2i at the time of CRTD implant, of these 55 patients started to assume these drugs; 4 and 2 patients already assumed ARNi and SGLT2i before CRT, respectively, and subsequently started the second drug. Thus, 100 patients were divided into groups according to ARNi and/or SGLT2i treatment after CRT.

In the supplemental material, Figure S5 summarizes the time of ARNi and SGLT2i start from CRT implant for each patient and Table S2 shows baseline characteristics and the average follow-up of the 2 groups.

Patients in the no ARNi/SGLT2i group were significantly older, more likely to have a low glomerular filtration rate.

The detailed distribution and changes in clinical and echocardiographic measurements are shown in Figures 4 and 5. NYHA functional class improved in 23 (37.7 %) patients and, importantly, decreased in only 2 (3.3 %) in ARNi/SGLT2i group vs 13 (33.3 %) in no ARNi/SGLT2i recipients (p < 0.001, Figure 4A). At mid-term follow-up 9 patients were in NYHA class I (mean LVEF 43.3 ± 5.12, all responders to CRT) with no indication to ARNi/SGLT2i prescription; all 4 patients in NYHA class IV (2 in ARNi/SGLT2i) had no clinical improvement. The highest benefits from ARNi/SGLT2i were found in NYHA class III where 19 (31 %) patients improved (Figure 4B). Of interest, ARNi and SGLT2i prescription had a similar impact on clinical improvement in both ICM (12, 35.3 %) and NICM (11, 40.7 %) patients (p = 0.75, Figure 4C). The ARNi/SGLT2i treatment improved significantly also the Δ LVEF, with a median increase of 4 % [IQR 2 %; 8 %] compared to the no treatment group -1.8 [0-2;-4] % (p < 0.001), without differences between ICM (Δ LVEF of 4 % [IQR 0.5-8 %]) and NICM (Δ LVEF of 4.5 % [IQR 2-6 %], p = 0.18, Figure 5A), nor with regard the mid-term LVEF (Figure 5B, p = 0.26). Figure 5C shows that patients in ARNi/SGLT2i group experienced consistent echocardiographic improvement: 23 (37.7 %) and 7 (11.5 %) of 5-9 % and > 10 % Δ LVEF increase, respectively.

**FIGURE 4.**
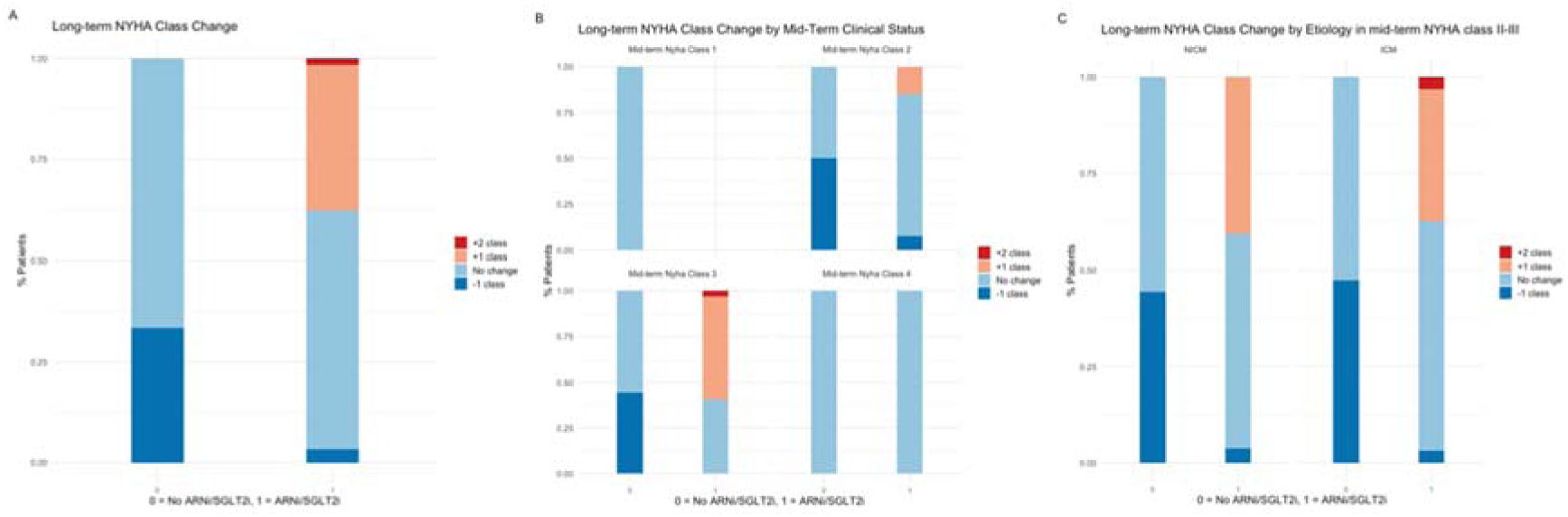
NYHA class change at long-term follow-up. ARNi/SGLT2i effectiveness on NYHA class change at long-term compared to mid-term follow-up in the entire population (N=100 patients with at least 24 months follow-up) **(A)**, divided according to mid-term NYHA class status **(B)** and to HF etiology **(C)**. At mid-term follow-up 9 patients were in NYHA class with no indication to ARNi/SGLT2i prescription; the highest benefits from ARNi/SGLT2i were found in NYHA class III. ARNi/SGLT2 prescription had a significant impact on clinical improvement regardless HF etiology (p value between ICM and NICM = 0.75). NICM refers to Non-Ischemic Cardiomyopathy, ICM to Ischemic Cardiomyopathy patients. Statistics: χ2 test.

**FIGURE 5.**
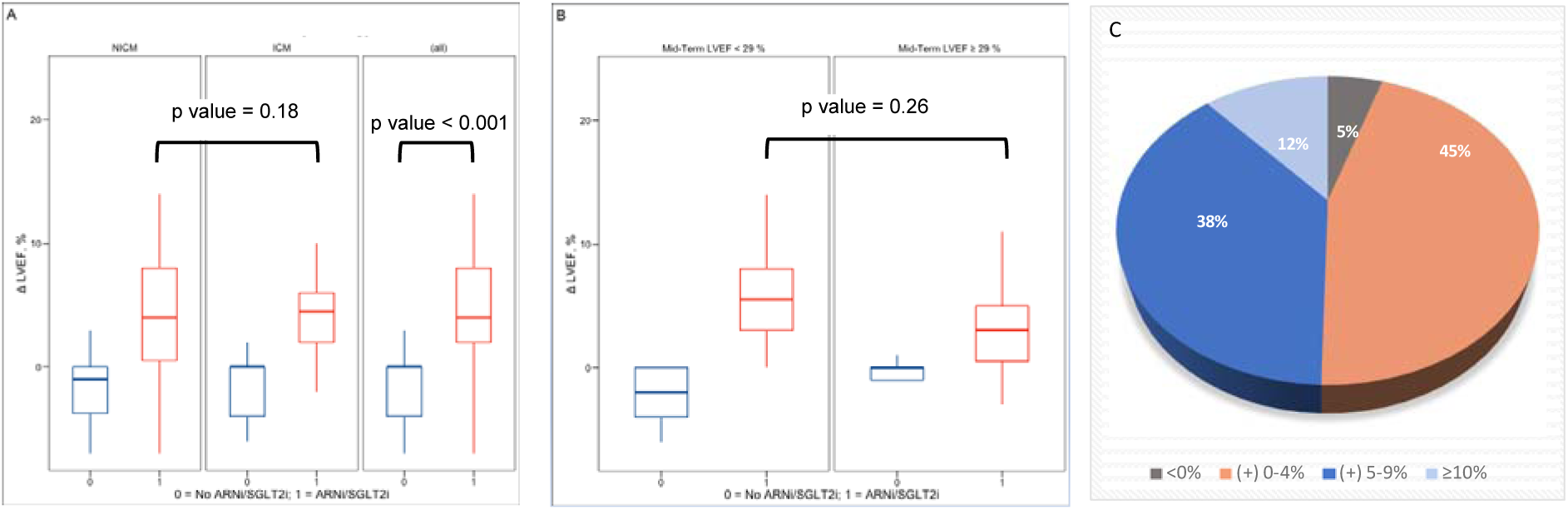
LVEF change in long-term analysis. Boxplots showing the ARNi/SGLT2i impact on Delta LVEF change at long-term compared to mid-term follow-up in all 100 patients and dividing by HF etiology **(A)** and mid-term median LVEF **(B)**. The advantages of ARNi/SGLT2i treatment were confirmed in both ICM and NICM patients and regardless to mid-term LVEF. In (B) patients were divided according to the median of mid-term LVEF (= 29%). The pie chart **(C)** shows the LVEF changes at long-term follow-up in the 61 ARNi/SGLT2i patients. 30 patients experienced consistent echocardiographic improvement: 23 of 5-9 % and 7 > 10 % Δ LVEF increase, respectively. NICM refers to Non-Ischemic Cardiomyopathy, ICM to Ischemic Cardiomyopathy patients. Statistics: Wilcoxon-Mann-Whitney test.

In addition, no significant differences have been found in clinical neither echocardiographic benefit dividing patients according to previous CRT response (See Figure S6 in the Supplement for details).

Finally, accounting the different time of mid-term to long-term follow-up between the 2 arms, Cox proportional hazards model shown in Table 4 assessed significant association of ARNi/SGLT2i initiation with the main clinical outcome of NYHA functional class II or I at long-term follow-up [Time-dependent ARNi/SGLT2i HR = 2.91, CI (1.23 – 6.9), p = 0.015]. After adjusting for confounders, ARNi/SGLT2i remained a strong positive predictor [Time-dependent ARNi/SGLT2i, HR = 3.67, CI (1.37, 10.2), p < 0.001]. Importantly, the graphic in Figure 6 plots the ARNi/SGLT2i estimate coefficient of the hazard ratio, resulting from a multivariable cox hazard proportional model adjusted for the same covariates as in the previous analysis but considering the entire period of follow-up: although the curve had a slight downward trend, the homogeneity of treatment effect according to the time period was respected [Time-dependent ARNi/SGLT2i, HR = 7.8 (3.4 – 14.71), Coefficient = 1.95, p for Schoenfeld residuals test = 0.102, Global Schoenfeld residuals test for the entire model, p = 0.158].

**FIGURE 6.**
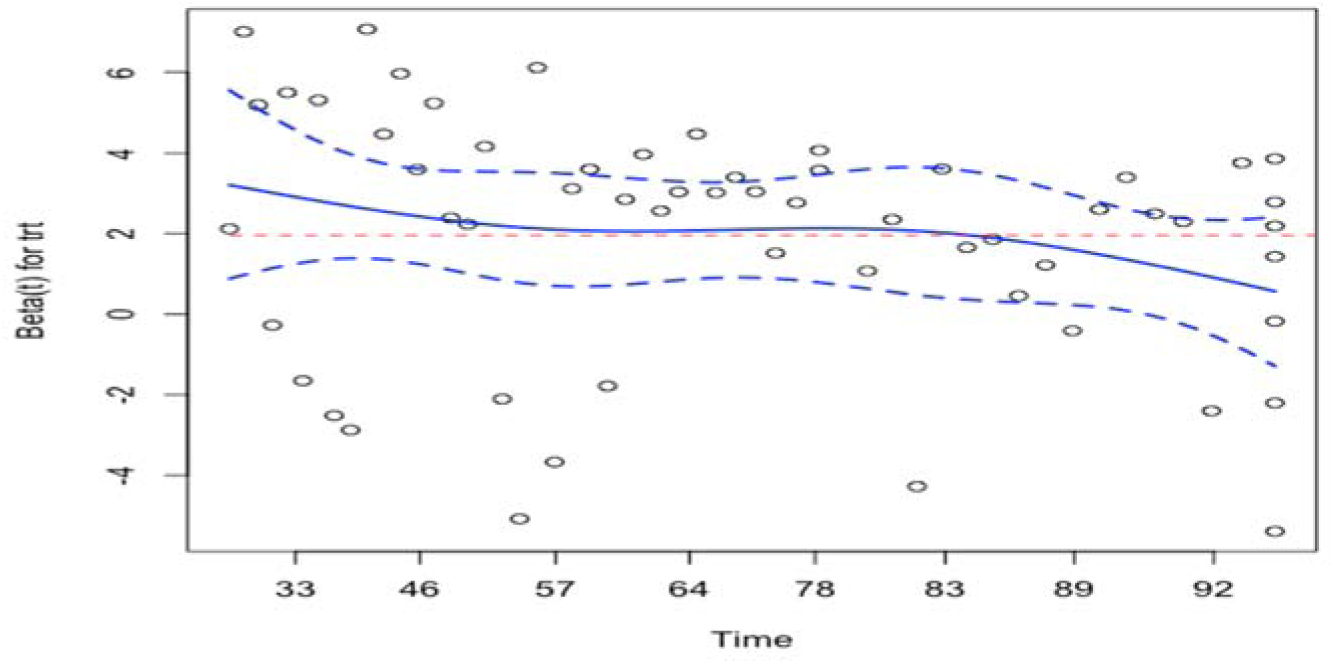
Effectiveness of ARNi and SGLT2i treatment in patients with long-term follow-up during the entire follow-up period. The plot shows proportional hazard assessment of ARNi/SGLT2i treatment in the multivariable Cox Regression analysis adjusted for the same covariates showed in Table 5 but accounting the entire follow-up period (from CRT implant). The dotted line in red refers to the coefficient of ARNi/SGLT2i time dependent covariate hazard ratio (plotted on the Y axis) derived from the model (Estimate coefficient = 1.95). The shape of the smoothed curve (in continuous blue line) is an estimate of the difference parameter as a function of time and appears to be constant through the time (in months, on the X axis), with a slightly decline only in the latter part. Dotted blue lines represent the 95 % confidence band for the smooth curve, the points the partial Schoenfeld residuals indicating the differences between the observed and the expected value of the covariate at each event time based on all those at risk at that time. Schoenfeld residuals test (p = 0.102) demonstrated that the proportional hazard assumption is satisfied and the effectiveness of ARNi/SGLT2i treatment is consistent through the entire period (Global Schoenfeld Residual test of the model, p = 0.158). “Trt” refers to ARNi and SGLT2i treatments.

**TABLE 4.** Factors associated with NYHA functional class I or II status at long-term compared to mid-term follow-up. Cox regression analysis in 100 long-term patients. In bold are reported significant values and the covariates entered in the multivariable analysis. ARNi and SGLT2i treatment was considered as time-dependent covariate. Linearity assumption for continuous covariates was tested with the Restricted cubic splines and ANOVA test in all continuous covariates and as for LVEF and LVESV resulted not respected (p = 0.02 and 0.031, respectively). In the multivariable model we have inserted LVEF due to a stronger association at univariable analysis compared to LVESV. * LVEF was included in multivariable model as restricted cubic spline to minimize residual confounding accounting the best fit of the model; consequently, Hazard Ratios and p values referring to the 4 different matrices of the transforming covariate, however not statistically significant, have not been reported. All the interactions exceeded 0.10 and Schoenfeld Residual tests resulted not significant (Global Schoenfeld Residual test, p = 0.212). Glomerular filtration rate was calculated in ml/min/1.73 m2. NICM = Non-Ischemic Cardiomyopathy; ICM = Ischemic Cardiomyopathy; COPD = Chronic Obstructive Pulmonary Disease; LVESV = Left Ventricular End-Systolic volume, in milliliters; LVEF = Left Ventricular Ejection Fraction.

| Variables | Hazard Ratios<br>(95 % CIs) | p value | Hazard Ratios<br>(95 % CIs) | p value |
| --- | --- | --- | --- | --- |
|  | Univariable analysis |  | Multivariable analysis |  |
| **ARNi/SGLT2i Treatment**<br>**(Time-dependent)** | 2.91 (1.23 – 6.9) | **0.015** | 3.67 (1.37 – 10.2) | **0.013** |
| Age (years) | 0.99 (0.97 – 1.03) | 0.92 | - | - |
| Atrial fibrillation at mid-term | 1.09 (0.76 – 1.55) | 0.93 | - | - |
| LVEF at mid-term, % | 1.07 (1.01 – 1.13) | **0.013** | ns* | ns* |
| LVESV at mid-term, ml | 0.95 (0.89 – 0.99) | **0.042** | - | - |
| NICM | Reference | na | - | - |
| ICM | 1.18 (0.66 – 2.11) | 0.572 | - | - |
| COPD | 0.55 (0.29 – 1.05) | 0.067 | 0.35 (0.17 – 0.75) | **0.006** |
| Glomerular filtration rate | 1.01 (0.99 – 1.02) | 0.28 | 1.01 (0.99 – 1.03) | 0.220 |
| Clinical CRT response | 1.88 (0.97 – 3.62) | 0.059 | 1.22 (0.57 – 2.61) | 0.591 |

**TABLE 5.**
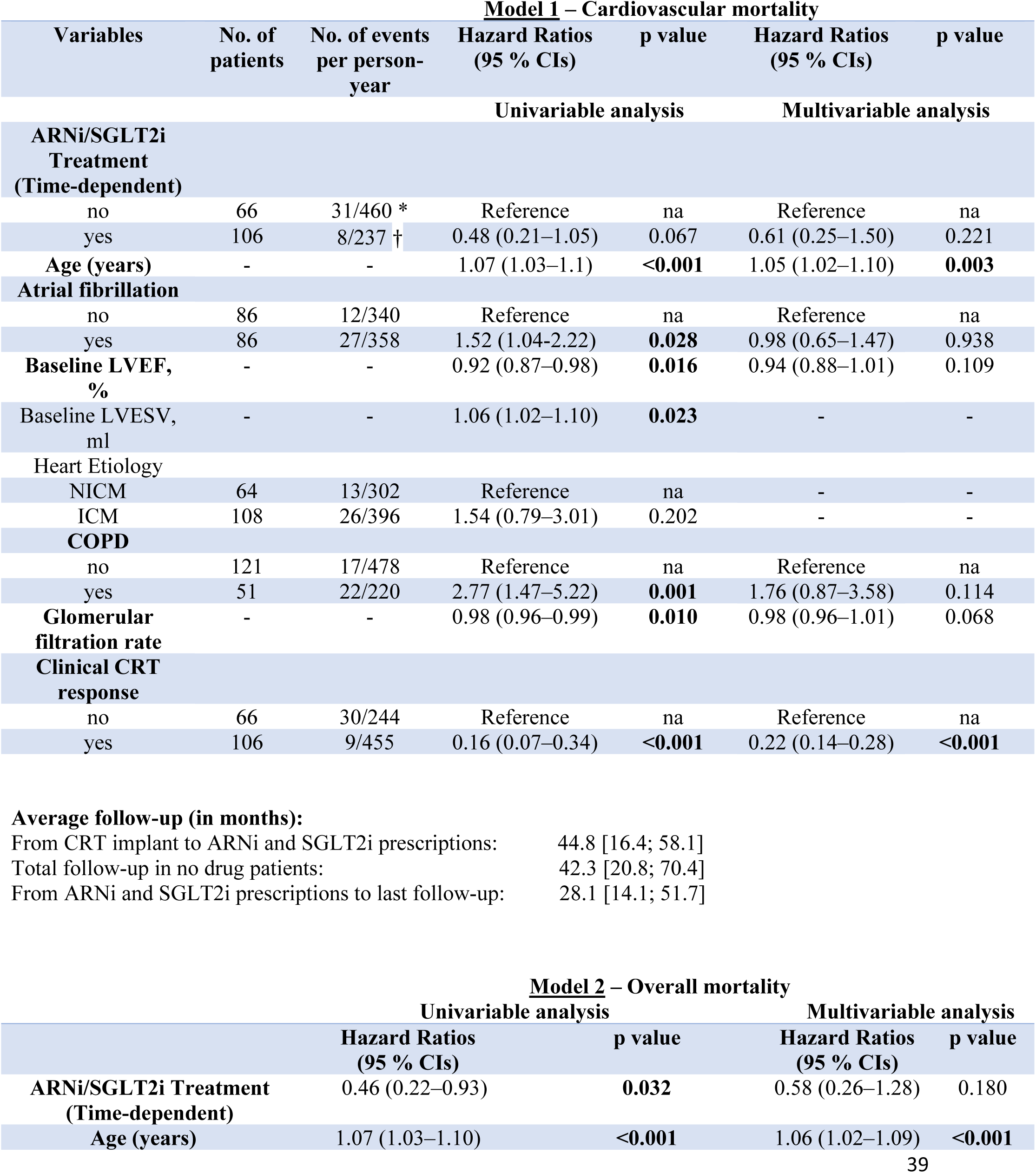
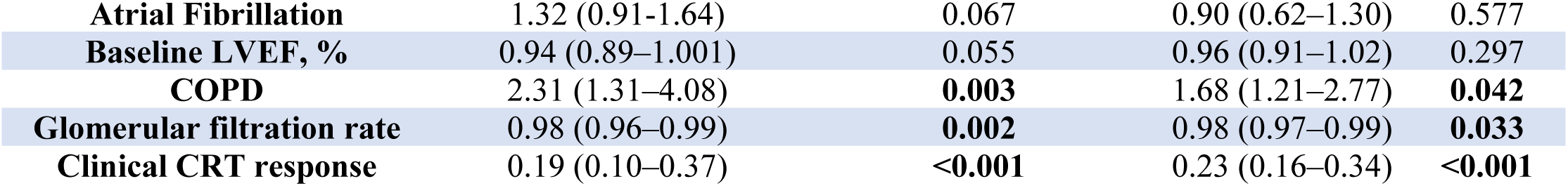
Factors associated with the occurrence of cardiovascular death and overall mortality according to univariable and multivariable Cox Regression analysis. Model 1 refers to factors associated with HF-related death, Model 2 to all-cause mortality. In model 2 are shown only covariates considered in the multivariable analysis. In bold are reported significant values and the covariates entered in the multivariable analysis. LVEF has been included in the multivariable models due to significant correlation and a stronger association at univariable analysis compared to LVESV. All the interactions with the main covariate were not significant. * 460 years were considered including the time of ARNi/SGLT2i patients follow-up before starting the treatments. † 237 years refer only to the period in ARNi/SGLT2i therapy. Glomerular filtration rate was calculated in ml/min/1.73 m2. NICM = Non-Ischemic Cardiomyopathy; ICM = Ischemic Cardiomyopathy; COPD = Chronic Obstructive Pulmonary Disease; LVESV = Left Ventricular End-Systolic volume, in milliliters; LVEF = Left Ventricular Ejection Fraction.

In Figure S7 (in the supplement) we described the main clinical and echocardiographic findings at long-term follow-up based on ARNi, SGLT2i or both treatments and ARNi doses. Once again, the effectiveness of S/V and gliflozins was confirmed in all subgroups analysis, but to a lesser degrees in the lowest S/V dose.

Plot in Figure 7 summarizes the main clinical findings of our study, showing the mean LVEF and the percentage of NYHA class I or II patients before CRT implant, at 12 months, mid- and long-term follow-up. Interestingly, pre-CRT ARNi/SGLT2i patients compared to post-CRT ARNi/SGLT2i and no treatments groups, displayed after 24 months from implant a greater clinical status (76.1 % in NYHA class I or II vs 69.1 % and 42,8 %) and a higher mean LVEF [36 ± 3 % vs 33.2 ± 2.9 (p = 0.042) and 30.1 ± 4.8 (p = 0.022), respectively]. Moreover, LVEF was also significant greater in post-CRT ARNi/SGLT2i vs no drugs patients (p = 0.048).

**FIGURE 7.**
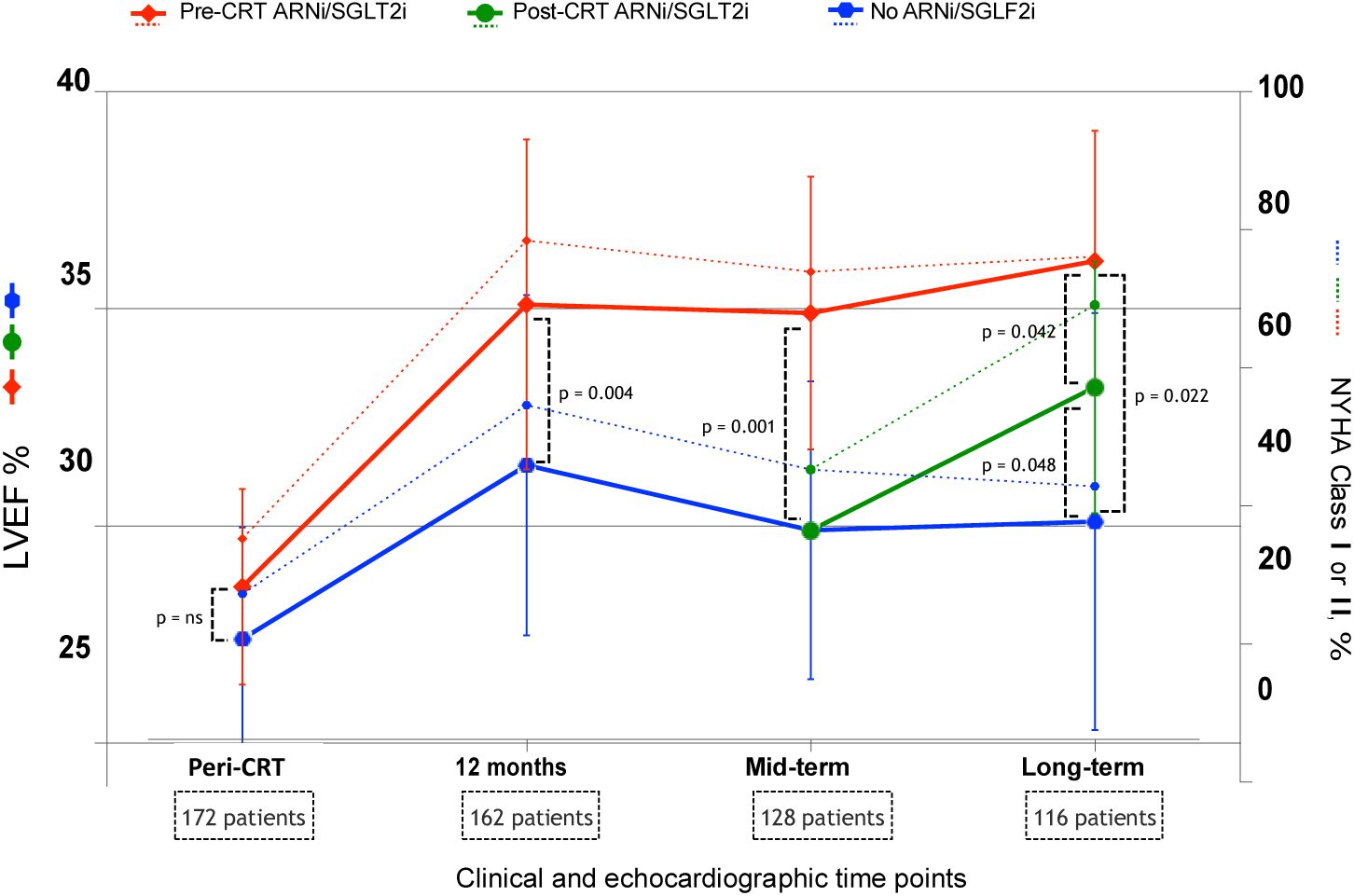
Median change of LVEF and the percentage of patients in NYHA class I or II. Plots showing the mean LVEF and the percentage of patients in NYHA class I or II before CRT implant, at 12 months, mid- and long-term follow-up in overall population study. Solid lines refer to LVEF (plotted on left Y axis), dotted lines to NYHA class I or II patients as percentage (plotted on right Y axis). For each follow-up points are reported the number of patients analyzed according to alive status and availability of long-term follow-up. Pre-CRT ARNi/SGLT2i patients displayed a greater LVEF and a better functional clinical status at 12-month follow-up and the benefit extended in all the assessments. ARNi and SGLT2i initiation at mid-term was associated again with a greater LVEF and a better functional clinical status at long-term compared to no drugs patients. At peri-CRT time point, no patients were in NYHA class I. Statistics: t-student test and pairwise test with the Holm correction for multiple comparison at long-term.

### Overall survival

During a median follow-up of 63.1 (CI 95 %, 52.7 - 73.8) months, 52 (30.2 %) patients died, 39 (22.7 %) due to cardiovascular HF-related death.

Kaplan-Meier estimates of HF and all-cause-death in the two study groups are shown in Figure 8. The curves diverged within the first 12 months and continue their separate paths thereafter (p < 0.001 in both survival functions). Nevertheless, patients who started the treatments in different time point have not been considered in this analysis.

**FIGURE 8.**
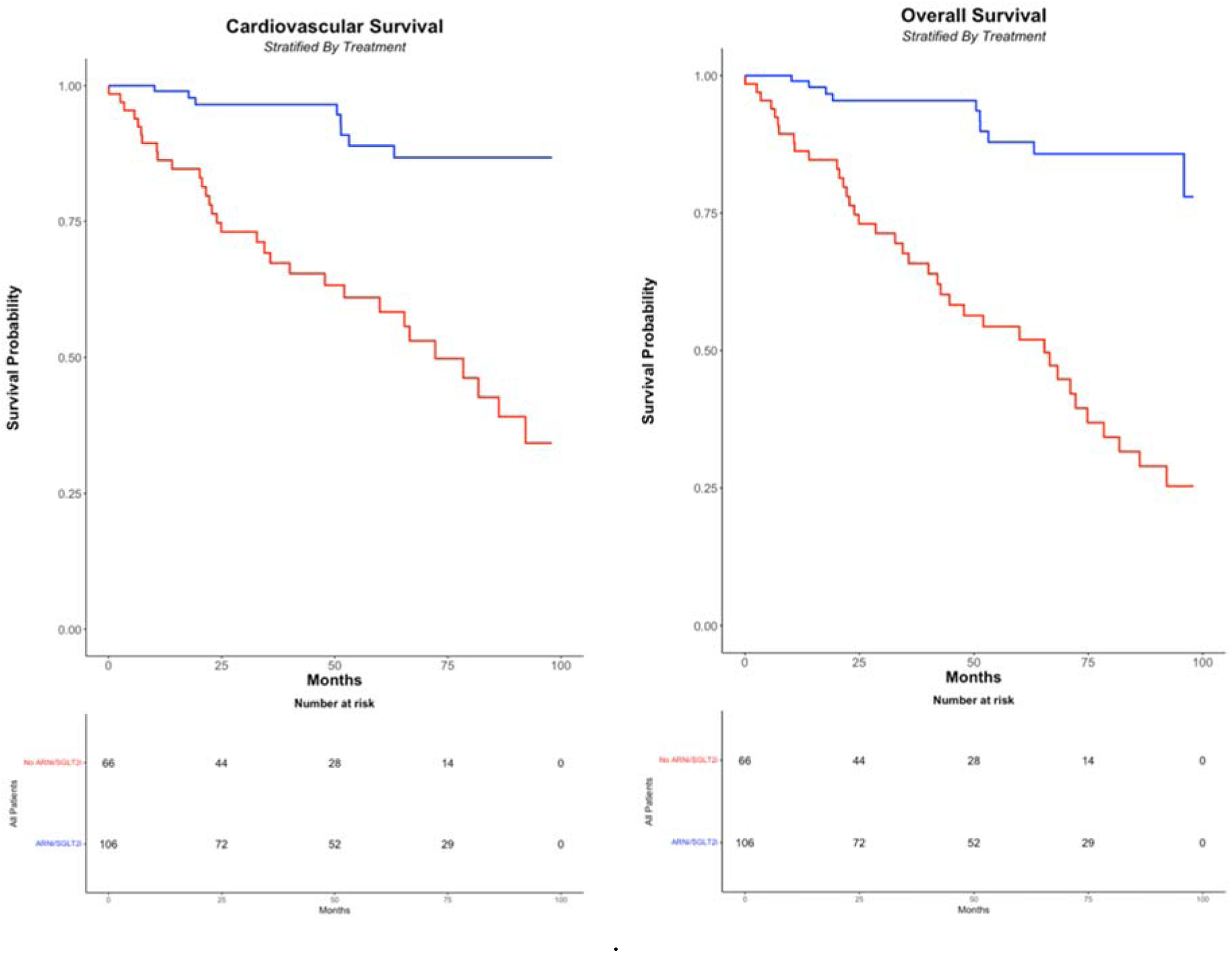
Kaplan-Meier estimates of the probability of survival free of heart failure and all-cause death. There were significant differences in the estimate of survival free of heart failure-related (graphic on the left) and all-cause death (graphic on the right) between the group that received ARNi or SGLT2i treatment during the entire follow-up and the no drugs patients (unadjusted p < 0.001 in both the analysis). Statistics: log-rank test.

In Table 5 are shown the number of cardiovascular deaths, the average follow-up times, and the hazard ratios for patients stratified according to the main clinical features. At univariable analysis, the effect size of ARNi/SGLT2i, considered as time-dependent variable, did not reach statistically significant reduction in HF death [p = 0.067, HR 0.48 (0.21 – 1.05)], confirmed after multivariable adjustment [p = 0.221, HR 0.61 (0.25 – 1.50)] (Table 5, model 1). Of note, clinical CRT response was the strongest predictors. Regarding overall mortality (Table 5, model 2), ARNi/SGLT2i treatment was a significant positive predictor only in univariable analysis [p = 0.032, HR 0.46 (0.22 – 0.93)] but not after adjusting for confounders [p = 0.180, HR 0.58 (0.26 – 1.28)].

## DISCUSSION

The main findings of our study are the following: 1) ARNi and SGLT2i increase clinical response rate to CRT and improve the cardiac function at 12-month follow-up; 2) their synergic benefits along with CRT are especially tangible in ICM patients; 3) an add-on ARNi and SGLT2i therapy is very effective on long-term echocardiographic and clinical status, regardless of HF etiology and of time of initiation from CRT implant; 4) their benefits on survival remain to be assessed, especially for the relatively limited follow-up period after ARNi and SGLT2i introduction in our study.

The CRT non-responsiveness constitutes a burning challenge, with around 30-50 % of patients not experiencing significant benefits ^20^; many efforts have been made for optimizing response, including the introduction of novel therapies. Among these, ARNi and SGLT2i have proven an overwhelming benefit in HF patients ^21^. However, in our best knowledge, no studies have evaluated their potential synergy with the contemporary CRT, and very few data have examined an add-on strategy with S/V and especially gliflozins administrated in CRT patients. Indeed, in the 2021 ESC guidelines for HF, ARNi and SGLT2i are a class I indication in patients NYHA class II-IV heart failure with LVEF ≤ 40 % to reduce the risk of HF hospitalization and death ^5^, but precise indications on a late combination with CRT are currently lacking.

### Impact of ARNi and SGLT2i in clinical and echocardiographic CRT response

To date in literature, little is known about the clinical and echocardiography impact of add-on ARNi/SGLT2i therapy among HFrEF patients with CRT during periprocedural period. The main trials that contributed to study the impact of ARNi/SGLT2i on outcomes in patients with HFrEF have only a little proportion of patients with CRT. As for the ARNi, in the PARADIGM-HF only 7% of the angiotensin-receptor-neprylisin inhibitor group had CRT ^2^, whereas in the PROVE-HF the 15,4% of the study group had it ^22^. As for the gliflozins, the DAPA-HF analyzed a population with only 8% of patients with CRT ^23^, whereas EMPEROR-Reduced with 11.8% ^24^. These trials didn’t analyze the impact of SGLT2i on these CRT patients.

In our cohort of 172 patients, approximately 75 % of ARNi and SGLT2i recipients experienced clinical and echocardiographic benefits 12-months after CRT implant and were classified as CRT responders. This rate was higher than in no ARNi/SGLT2i group, however it reached statistical significance only in ICM patients. Previous studies reported that CRT reduced all-cause mortality similarly in both ICM and NICM patients ^25^, however sub-analysis of randomized studies demonstrated the occurrence of more favorable reverse remodeling in NICM compared to ICM ^26–29^. In the REVERSE study, 50-59 % in ICM vs 74-83 % in NICM group (based on different response criteria) were considered CRT responders after 1-year follow-up ^27^, percentages similar to those of our no ARNi/SGLT2i group. By contrast, in our study the effectiveness of CRT in ARNi and SGLT2i group was consistent regardless HF etiology, but especially in ICM patients that least responded to the CRT. Interestingly, the other well-known negatively predictors of CRT clinical outcomes (such as atrial fibrillation, RBBB, biventricular pacing) were confirmed in our analysis ^30–32^.

Considering only echocardiographic parameters, our data proved that the effect of ARNi and SGLT2i was consistent in both ICM and NICM patients, with a greater Δ LVEF increase compared to only CRT group. In a meta-analysis of over 10.000 patients, S/V was associated with a mean LVEF increase of +5,11 % compared to patients treated with ACEI/ARB therapy and a similar effect was confirmed in the prospective PROVE-HF study ^22, 33^. Regarding SGLT2i, likewise the two recent SUGAR-DM-HF and EMPA-TROPISM trials suggested favorable reverse remodeling in term of LVESV reduction and both LVESV reduction and LVEF improvement, respectively ^34, 35^. Our data are concordant with these literature findings and assume a potential synergistic role of S/V and gliflozins in addition to CRT.

### ARNi and SGLT2i effectiveness in long-term follow-up

Based on current evidence, about 40 % of CRT recipients are potentially indicated for medical therapy optimization with S/V and gliflozins ^9, 36, 37^. Furthermore, it is essential to determine the effectiveness of ARNi and SGLT2i in patients with CIED and in particular CRT.

The TAROT-HF study demonstrated that the effectiveness of S/V was greater in non-CRT-eligible patients based on QRS morphology; in fact, the improvements of LVEF and LVESV were more significant in this group compared to bundle branch block and a QRS > 130 msec^38^.

In a recent study, Russo et al. analyzed the impact of ARNi in190 CRTD non-responder patients, with a median follow-up of 20 months from device implant; about 20 % of their population improved cardiac function and were classified as additional responders ^39^.

In our analysis, ARNi and SGLT2i initiation after CRT implant was associated with improvement of cardiac function and a probability about 3-fold higher to be in class NYHA I or II at long-term follow-up, compared to no treatment patients, and only few patients experienced clinical deterioration. In addition, our population had a long follow-up, and the benefits of the treatment were confirmed along the entire period. Although CRT is an essential treatment for heart failure at any stage, HF is a progressive disease, and it does not cure the underlying disease. Considering the natural course of disease, many patients initially responders could clinically deteriorate. In our population, the effectiveness of ARNi and SGLT2i was confirmed both in responder and non-responder patients, and many patients initially responders have worsened their clinical status at long-term follow-up, or despite an improvement after CRT implant, remained symptomatic and needed medical optimization.

Finally, there is a lack of data about gliflozins effectiveness in these setting of patients. Although in our study we considered ARNi and SGLT2i together for analysis, from our data it appears that both had a significant impact in clinical and echocardiographic function. Further studies have to confirm the role of gliflozins in CRT recipients.

### ARNi and SGLT2 impact on survival

Sacubitril/valsartan and gliflozins were shown to reduce mortality and hospitalization rate in HFrEF patients. An indirect comparison of ARNi vs placebo using data from PARADIGM-HF and CHARM-Alternative studies demonstrated a 48 % reduction in HF death and hospitalization in S/V group ^40^. Among 8474 patients enrolled in DAPA-HF and EMPEROR-Reduced trial, the estimated gliflozins treatment effect was a significant 26 % relative reduction in the combined cardiovascular death and first HF hospitalization ^7^. Regarding patients with cardiac implantable electronic devices, a post hoc analysis of PARADIGM-HF analyzed the subgroup of patients with ICD or CRT-D and S/V was confirmed superior to enalapril regarding the primary endpoint of a composite of HF death and hospitalization ^8^. Moreover, a recent sub-analysis of DAPA-HF showed no significant interaction between gliflozins and CRT ^41^. In addition, S/V significantly reduced Sudden cardiac death (SCD) in patients eligible to implantable defibrillator therapy but without ICD ^42^, but this effect was more pronounced after the first months of therapy ^43^. The effectiveness of S/V on SCD reduction was confirmed in a meta-analysis of Liu et al. ^44^. These findings suggested that the underlying mechanism for the prevention of SCD is different from that of an ICD, probably related to the reverse remodeling. Indeed, Martens et al. confirmed that in 110 HFrEF patients with ICD or CRT, those who manifested an improvement in LVEF had a more pronounced reduction in premature ventricular contractions and non-sustained ventricular tachycardia burden ^45^. Regarding gliflozins, in a further post hoc analysis of DAPA-HF, during a median follow-up of 18 months, dapagliflozin reduced the risk of the primary endpoint of any serious ventricular arrhythmia, cardiac arrest or SCD and the effectiveness appeared to be more substantial in patients without a defibrillating device, although the interaction with dapagliflozin was not significant; as authors discussed, the device subgroup was of modest size with relatively few events, but above all ICD and CRT recipients were considered together not allowing to evaluate the possible synergic effect of CRT and dapagliflozin on cardiac remodeling and life-threatening arrhythmic events ^46^. Conversely, a meta-analysis of Sfairopoulos et al. failed to demonstrate a significant association of SGLT2i and the risk of SCD or ventricular arrhythmias ^47^. Further research is needed to better elucidate the effectiveness of these drugs on SCD reduction and the possible different underlying mechanism between ARNi and SGLT2i regarding their benefits in HF patients. As for only CRT patients, in a retrospective study of 50 patients who were CRT non responders, S/V treatment was associated with a lower risk of cardiac death, heart transplantation and left ventricular assist device (p = 0.029 using Kaplan-Meier method)^48^. In our survival analysis, Kaplan-Meier curves showed a similar trend in the estimate of survival free of heart failure according to treatment. Nevertheless, S/V and gliflozins are time dependent covariates: patients who received the treatments had to live long enough to receive those treatments, also it is worthwhile to consider that most patients could not take these drugs during the first follow-up years when their benefits were not yet well known and before the 2021 ESC HF recommendation. Accordingly, in the analysis when considering ARNi and SGLT2i as time dependent covariates, their effect size has not been shown to reduce the HF-related death risk in the framework of Cox proportional hazards models. On the other hand, the follow-up after their introduction was considerably shorter than the entire period of patients without or before starting the treatments and this may have limited the power of our analysis. Further studies are needed to assess the impact of S/V and gliflozins on survival in this setting of patients.

This study has several limitations, that are the following. First, the retrospective nature of our study made the data collection and complete retrieval of information challenging.

Furthermore, we relied on hospital records and/or on trans-telephonic interviews with patients or caregivers to determine mortality.

Second, we don’t have data regarding 6-minute walk test, B-type natriuretic peptide and N-terminal pro B-type natriuretic peptide, and all of these may be a surrogate of reverse remodeling as shown in previous studies.

Third, the clinical and echocardiographic changes seen after 12 months from CRT implant were attributed exclusively to the assumption of ARNi/SGLT2i, however several studies in literature have demonstrated that a late-onset response to CRT is possible ^49, 50^. However, in these studies, post-CRT improvement is greater in first-year post-implantation and mainly far beyond 60 months which corresponds to our median follow-up.

Fourth, our analysis has the weakness to consider both gliflozins and S/V in the analysis, due to the low sample after dividing for subgroups: thus, we only briefly described the individual effectiveness.

Finally, although our study had a relatively small sample size, to our knowledge this is the first study analyzing clinical and echocardiographic CRT response in patients with ARNi and SGLT2i, and we also provided a long-term follow-up to analyze their effectiveness after CRT implant. Therefore, our study fills a gap in current knowledge and may, despite its limitations, encourage future prospective well sampled studies to assess the synergistic impact of ARNi and SGLT2i (also considered individually).

## CONCLUSION

The ARNi and SGLT2i treatment in CRT patients improves the clinical and echocardiographic response at 12 months follow-up and at long-term follow-up, independently from the time of initiation. These drugs also have benefit on survival, however further studies are needed to confirm these data.

## Non-standard Abbreviation and Acronyms

ARNi: Angiotensin receptor-neprilysin inhibitor
COPD: Chronic obstructive pulmonary disease
CRT: Cardiac resynchronization therapy
HF: Heart failure
HFrEF: Heart failure and reduced ejection fraction
ICM: Ischemic cardiomyopathy
LVEF: Left ventricular ejection fraction
LVESV: Left ventricular end-systolic volume
NICM: Non-ischemic cardiomyopathy
SGLT2i: Sodium-glucose co-transporter 2 inhibitors

## Data Availability

The data that support the findings of this study are available from the corresponding author upon reasonable request. The corresponding author has full access to all the data in the study and takes responsibility for its integrity and the data analysis.

## ACKNOWLEDGMENTS

None.

## Supplemental Material

Tables S1-S2

Figures S1-S7

